# Longevity of SARS-CoV-2 Antibody in Health Care Workers: 6-Months Follow Up

**DOI:** 10.1101/2021.08.25.21262632

**Authors:** Michael Brant-Zawadzki, Deborah Fridman, Philip A. Robinson, Randy German, Arell Shapiro, Marcus Breit, Stacy Wilton, Elmira Burke, Jason R. Bock, Junko Hara

## Abstract

The prevalence and longevity of acquired immunity to coronavirus disease 2019 (**COVID-19**) in health care workers (**HCWs**) is of great interest, especially with the roll-out of vaccines for SARS-CoV-2. Determining such immunity may enhance knowledge about susceptibility of HCWs to COVID-19, frequency of vaccine administration, and degree of workplace risk, and may also support enactment of better workplace policies and procedures.

The present study reports on 6-months follow-up serosurveillance to determine the longevity of SARS-CoV-2 antibodies in HCWs.

Sub-sample (n=35) of the original serosurveillance in HCWs (n = 3,458) with baseline, 8-week, and 6-month blood sampling were analyzed. Information on job duties, location, COVID-19 symptoms, polymerase chain reaction test history, travel since January 2020, and household contacts with COVID-19 was collected.

Of 35 subjects, 13 were seropositive at baseline and maintained positivity at 8-week follow-up, with 3 losing positivity at 6-month follow-up. Among 22 subjects who were seronegative at baseline and seropositive at 8-week follow-up, all but one maintained positivity at 6-month follow-up. There was no significant effect of all factors (e.g., age, gender, job duties) examined at the .05 level on seropositivity at 6-month follow-up. The observed antibody longevity was 7.0+/-0.6 months for seropositive subjects (n=13), and 4.5+/-0.8 months for those seronegative subjects (n=22), at baseline. The longest duration of seropositivity observed in this cohort was 7.9 months (236 days).

With reported COVID-19-related symptoms up to 4.7 months prior to baseline blood sampling, possibly longer antibody presence is suggested. Similarly, seropositivity at 6-month follow-up further suggests greater antibody longevity than observed in this study.

## Introduction

Prior sero-suvaillance studies among HCWs have reported widely ranging antibody prevalence from 89.3% in Wuhan, China (n=424) [1], 35.8% in New York City (n=285) [2], to 7.4% in Milan, Italy [3], and 2.67% in Denmark (n=28,792) [4]. Our previous HCW sero-suvaillance studies have shown a low prevalence of antibodies ranging from 0.98% to 2.58% depending on sampling timing (e.g., before or after partial state re-opening in late May, 2020) [5,6]. Serology prevalence in HCWs was found to be loosely correlated with community prevalence [6].

The longevity of antibody presence has also been reported, though sparsely, with varying range among HCWs [7-9]. This study reports 6-month follow-up serosurveillance conducted on sub-samples from our HCW cohort studied in May through August, 2020, at Hoag Memorial Hospital Presbyterian, California, United States.

## Methods

### Subject Recruitment, Enrollment, and Data Collection

IRB approval was obtained for this study (Providence St. Joseph Health IRB # 2020000337). The original HCWs cohort (n=3,458) was recruited by email notification to the entire employee workforce and medical staff [6]. Consenting participants were interviewed with regards to job duties, location, COVID-19 symptoms (i.e., fever, sore throat, cough, rhinorrhea, loss of taste or smell), PCR test history, travel record, and existence of household contacts with COVID-19. Job duties were used to further classify subjects into high (e.g., MD, RN, ICU tech), medium (e.g., phlebotomist, medical tech), or low (e.g., admin, IT) risk groups to stratify levels of direct exposure to COVID-19. A blood sample (∼5ml) was collected at enrollment and 8-week follow-up, from each subject for serum analysis for IgG antibodies to SARS-CoV-2 using the VITROS Anti-SARS-CoV-2 IgG Reagent Pack and Calibrator on the VITROS® XT 7600 instrument by Ortho Clinical Diagnostics approved for the FDA emergency use authorization.

### 6-Month Blood Sampling

Additional blood samples were collected at 6-month follow-up from those who had positive antibody results at the initial and 8-week follow-up, or at 8-week follow-up only, with a total of 96 subjects eligible. However, due to the COVID-19 vaccine roll-out around the time of 6-month follow-up, with some subjects already vaccinated by the time of their blood sampling, 61 subjects were excluded from analysis, resulting in 35 subjects (36%) included in this study. The study was discontinued after January 24, 2021.

#### Analysis

A Mann-Whitney U test was used to assess the significance of age, and a series of one-sided Fisher’s exact tests were used for gender, race, and job risk level, on seropositivity at 6-month follow-up. Longevity of antibody positivity was plotted for each subject along with a self-reported polymerase chain reaction (**PCR**) test history, onset of COVID-19 related symptoms, prior travel record, close contract with COVID-19 patients, and rank-ordered by baseline antibody results, and then job risk levels.

## Results

**Table 1** summarizes sample characteristics (n=35). Of 35 subjects, 13 were seropositive at baseline and maintained positivity at 8-week follow-up, with 3 losing positivity at 6-month follow-up. Among 22 subjects who were seronegative at baseline and seropositive at 8-week follow-up, all but one maintained positivity at 6-month follow-up. A Mann-Whitney U test and Fisher’s exact tests showed no significant effect of all factors examined at the .05 level on seropositivity at 6-month follow-up.

**Table 1.** Sample Characteristics.

|  | Job Low Risk | Job Medium Risk | Job High Risk | Total |
| --- | --- | --- | --- | --- |
|  | n = 9 (26%) | n = 4 (11%) | n = 22 (63%) | N = 35 |
| Baseline - Antibody Positive, count (%) | 4 (44%) | 3 (75%) | 6 (27%) | 13 (37%) |
| 2-month - Antibody Positive, count (%) | 9 (100%) | 4 (100%) | 22 (100%) | 35 (100%) |
| 6-month - Antibody Positive, count (%) | 7 (78%) | 4 (100%) | 20 (91%) | 31 (89%) |
| Age in yrs., *M (SD)* | 35.44 (8.83) | 36.25 (4.57) | 34.05 (9.42) | 34.66 (8.70) |
| Female, count (%) | 7 (78%) | 2 (50%) | 19 (86%) | 28 (80%) |
| Race, count (%) |  |  |  |  |
| American Indian or Alaska Native | 0 | 0 | 0 | 0 |
| Asian | 2 (22%) | 1 (25%) | 5 (23%) | 8 (23%) |
| Black | 0 | 0 | 0 | 0 |
| Hispanic or Latino | 3 (33%) | 2 (50%) | 3 (14%) | 8 (23%) |
| Native Hawaiian or Pacific Islander | 1 (11%) | 0 | 1 (5%) | 2 (6%) |
| White | 3 (33%) | 1 (25%) | 11 (50%) | 15 (43%) |
| Other | 0 | 0 | 2 (9%) | 2 (6%) |

**Figure 1** shows longevity of antibody positivity, demonstrating 7.0+/-0.6 months of longevity for seropositive subjects (n=13), and 4.5+/-0.8 months for those seronegative subjects (n=22), at baseline. The longest duration of seropositivity observed in this cohort was 7.9 months (236 days).

**Figure 1.**
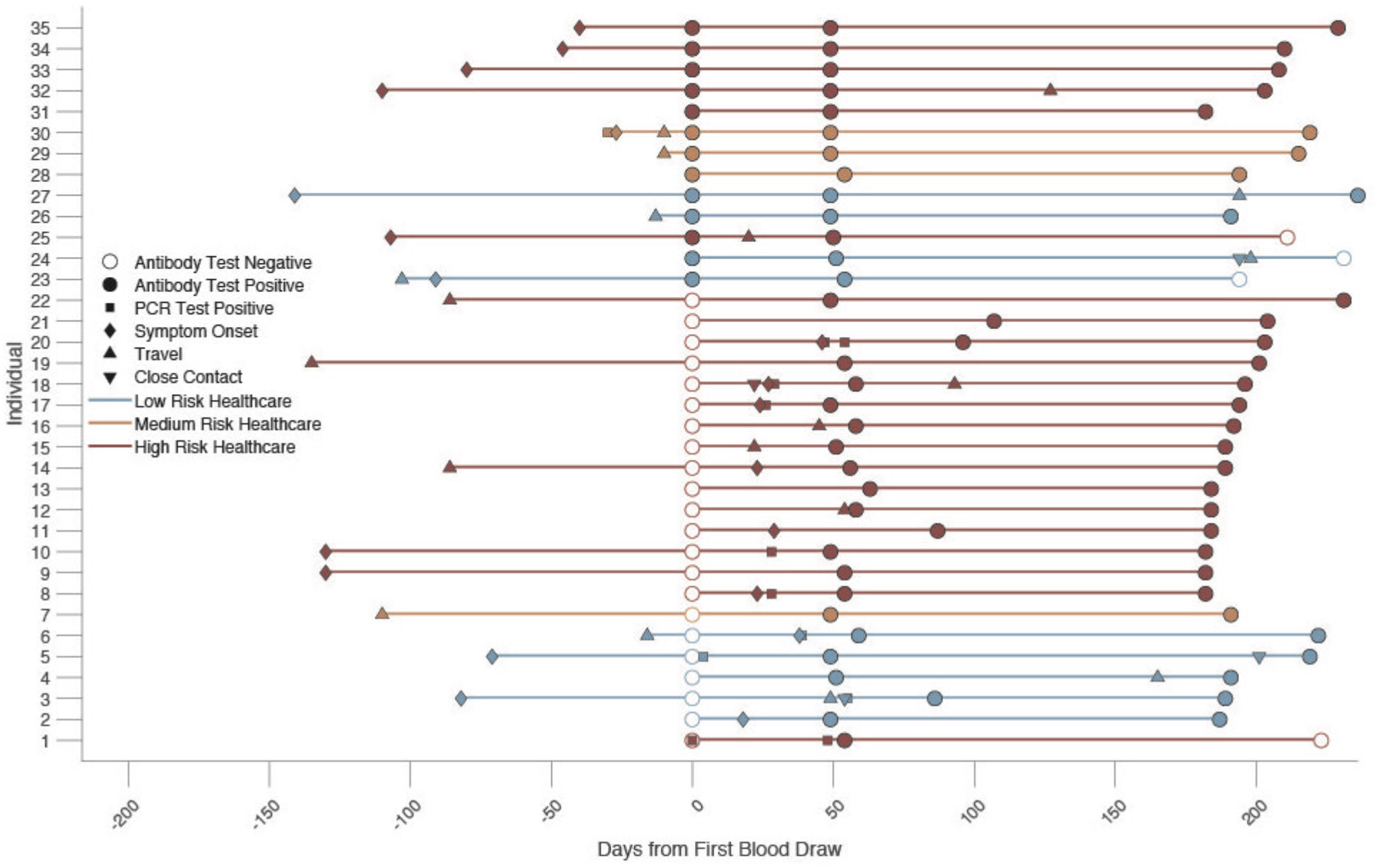
SARS-CoV-2 antibody positivity. SARS-CoV-2 antibody positivity at baseline (0 at x-axis), 8-week, and 6-month follow up are plotted for each subject. Other factors possibly affecting antibody positivity including self-reported COVID-19 diagnosis, COVID-19 symptoms onset, and travel history are also included.

## Discussion

Our data suggests antibody longevity of up to 7.9 months, with possibly greater longevity, in our HCWs. This finding corroborates other antibody longevity studies reporting greater longevity than previously observed in mixed cohort of patients, HCWs, and general public [10-12], up to 8 months of longevity [11].

Among those seropositive at baseline, eight reported COVID-19-related symptoms up to 4.7 months prior to baseline blood sampling, suggesting possibly longer antibody presence. Similarly, the fact that all seropositive cases except four maintained seropositvity at 6 months, further suggests greater antibody longevity than observed in this study.

This finding is encouraging for both those with acquired immunity from subclinical infection, and those who would acquire immunity by vaccination. A study from the National Cancer Institute reports that people with positive antibodies have substantial immunity to SARS-CoV-2, which means they may be at lower risk for future infection [13]. It is not known whether waning antibody presence equates to a loss of T-cell based immunity. Our findings may help guide prioritization of HCWs for immunization based on their serology history.

## Data Availability

All the data referred in this study will be available through figshre.com.

## Acknowledgement

We acknowledge our healthcare workers who have contributed to this study.

